# A Multimodality Video-Based AI Biomarker For Aortic Stenosis Development And Progression

**DOI:** 10.1101/2023.09.28.23296234

**Authors:** Evangelos K. Oikonomou, Gregory Holste, Neal Yuan, Andreas Coppi, Robert L. McNamara, Norrisa Haynes, Amit N. Vora, Eric J. Velazquez, Fan Li, Venu Menon, Samir R. Kapadia, Thomas M Gill, Girish N. Nadkarni, Harlan M. Krumholz, Zhangyang Wang, David Ouyang, Rohan Khera

## Abstract

**Importance:** Aortic stenosis (AS) is a major public health challenge with a growing therapeutic landscape, but current biomarkers do not inform personalized screening and follow-up.

**Objective:** A video-based artificial intelligence (AI) biomarker (Digital AS Severity index [DASSi]) can detect severe AS using single-view long-axis echocardiography without Doppler. Here, we deploy DASSi to patients with no or mild/moderate AS at baseline to identify AS development and progression.

**Design, Setting, and Participants:** We defined two cohorts of patients without severe AS undergoing echocardiography in the Yale-New Haven Health System (YNHHS) (2015-2021, 4.1[IQR:2.4-5.4] follow-up years) and Cedars-Sinai Medical Center (CSMC) (2018-2019, 3.4[IQR:2.8-3.9] follow-up years). We further developed a novel computational pipeline for the cross-modality translation of DASSi into cardiac magnetic resonance (CMR) imaging in the UK Biobank (2.5[IQR:1.6-3.9] follow-up years). Analyses were performed between August 2023-February 2024.

**Exposure:** DASSi (range: 0-1) derived from AI applied to echocardiography and CMR videos.

**Main Outcomes and Measures:** Annualized change in peak aortic valve velocity (AV-V_max_) and late (>6 months) aortic valve replacement (AVR).

**Results:** A total of 12,599 participants were included in the echocardiographic study (YNHHS: *n*=8,798, median age of 71 [IQR (interquartile range):60-80] years, 4250 [48.3%] women, and CSMC: *n*=3,801, 67 [IQR:54-78] years, 1685 [44.3%] women). Higher baseline DASSi was associated with faster progression in AV-V_max_ (per 0.1 DASSi increments: YNHHS: +0.033 m/s/year [95%CI:0.028-0.038], n=5,483, and CSMC: +0.082 m/s/year [0.053-0.111], n=1,292), with levels ≥ vs <0.2 linked to a 4-to-5-fold higher AVR risk (715 events in YNHHS; adj.HR 4.97 [95%CI: 2.71-5.82], 56 events in CSMC: 4.04 [0.92-17.7]), independent of age, sex, ethnicity/race, ejection fraction and AV-V_max_. This was reproduced across 45,474 participants (median age 65 [IQR:59-71] years, 23,559 [51.8%] women) undergoing CMR in the UK Biobank (adj.HR 11.4 [95%CI:2.56-50.60] for DASSi ≥vs<0.2). Saliency maps and phenome-wide association studies supported links with traditional cardiovascular risk factors and diastolic dysfunction.

**Conclusions and Relevance:** In this cohort study of patients without severe AS undergoing echocardiography or CMR imaging, a new AI-based video biomarker is independently associated with AS development and progression, enabling opportunistic risk stratification across cardiovascular imaging modalities as well as potential application on handheld devices.

## INTRODUCTION

With the expanding availability of transcatheter and surgical aortic valve replacement (AVR) procedures that effectively modify the prognosis of aortic stenosis (AS),^1–4^ there has been a focus on the timely identification of individuals at risk of rapid progression.^5–8^ Such efforts have been limited by the high prevalence of aortic sclerosis, variable progression rates, ^9–11^ and marked heterogeneity in the condition and its drivers.^12–15^ As monitoring continues to depend on referrals for comprehensive testing by Doppler echocardiography, there is an unmet need to develop more precise algorithms to better define the personalized trajectory of AS.

We recently developed a deep learning (DL) strategy that uses a self-supervised, contrastive learning approach to learn the computational representation of severe AS on simple parasternal long axis (PLAX) videos without Doppler,^16^ a standard echocardiographic view easily captured on handheld devices.^16,17^ The predicted model-derived phenotype score, the Digital AS Severity index (DASSi), demonstrated excellent performance (Area Under the Receiver Operating Characteristic curve [AUROC] of 0.94-0.98) for the discrimination of severe AS across 9,338 studies in three distinct cohorts.^16^

In the present study, we hypothesized that, as DASSi identifies the echocardiographic signature of severe AS, it may stratify the risk of AS development and progression among individuals without AS or with early valve sclerosis and stenosis, independent of traditional Doppler parameters. Furthermore, we hypothesized that the anatomical and temporal information learned by DASSi through self-supervised pre-training on a video-based modality^16^ would generalize to other modalities where the cinematic representation of cardiac activity is captured. We examined this using data from two health system-based echocardiography cohorts across the U.S., as well as cardiac magnetic resonance (CMR) data from the UK Biobank^18^ after developing a computational pipeline for cross-modal translation, thus spanning a range of geographical settings, imaging modalities, and phenotypes.

## METHODS

### Study Design, Study Population and Data Source

#### Echocardiography Study

This was a multi-center retrospective cohort study of patients without severe AS (with moderate, mild, or no AS) who underwent clinical echocardiography and were followed longitudinally within their respective health systems (**Figure 1A**). Participants were independently drawn from 5 hospitals affiliated with Yale-New Haven Health System (YNHHS) across Connecticut and Rhode Island between 2015-2021, and from Cedars-Sinai Medical Center (CSMC) (Los Angeles, California) between 2018-2019 to define two nested sub-cohorts within each center: (1) A *<u>longitudinal echocardiography cohort</u>* of individuals undergoing clinically-indicated transthoracic echocardiography (TTE) at two or more timepoints; and (2) a *<u>clinical</u> <u>outcomes cohort</u>* which included all individuals with longitudinal follow-up for AVR. Eligible individuals had i) baseline peak aortic valve velocity (AV-V_max_) of <4 m/sec, ii) no prior AVR, and iii) PLAX videos available for processing. To avoid bias, none of the patients from the original training set were included.^16^ Moreover, the YNHHS cohort was specifically enriched for cases of mild/moderate AS, whereas the CSMC cohort sampled cases at random, reflecting local AS prevalence (***Supplement***).

**Figure 1.**
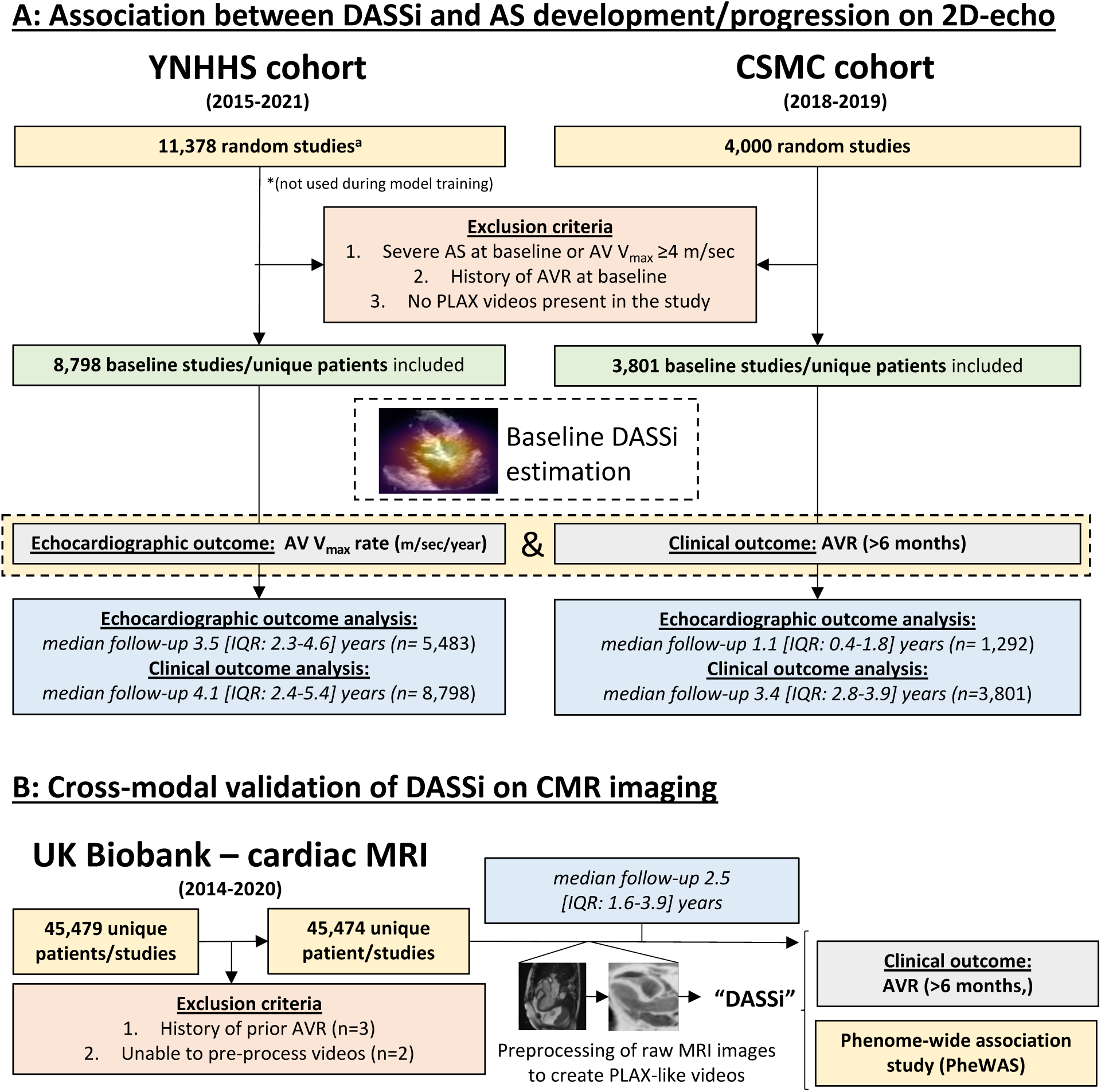
Study design. AV-V_max_: peak aortic valve velocity; AVR: aortic valve replacement; CMR: cardiac magnetic resonance; DASSi: digital aortic stenosis severity index; PLAX: parasternal long axis view.

#### CMR study/UK Biobank

The UK Biobank represents a prospective observational study of 502,468 participants aged 40-69 years. We performed a post-hoc analysis on 45,474 individuals (182 [0.4%] patients with AS of undetermined severity by diagnosis codes) who enrolled in the CMR sub-study between 2014-2020,^18,19^ after excluding individuals who had withdrawn consent, had prior AVR or videos that failed technical preprocessing (**Figure 1B**).

Across all three cohorts reporting stands consistent with the STROBE statement.^29^ Race and ethnicity were self-reported by the participants. In the YNHHS cohort, race categories included African American, Asian, White, and other, which included American Indian or Alaskan Native, Native Hawaiian or Pacific Islander, more than one race, unknown, or not reported. Hispanic ethnicity was reported separately. In the CSMC cohort, Hispanic ethnicity was included as an additional field under race/ethnicity. In the UKB, race groups included Asian (or Asian British, or Chinese), Black (or Black British), White, and other, which included more than one race, unknown, or not reported.

#### Echocardiogram/CMR Interpretation

All clinical echocardiograms were reviewed by board-certified cardiologists in the local laboratories who determined the presence and severity of AS according to the American Society of Echocardiography.^9,20^ Measurements other than DASSi were derived from the structured echocardiographic reports. The CMR studies in the UK Biobank were performed as part of a research protocol, with left ventricular ejection fraction (LVEF) measurements previously measured in an automated manner (***Supplement***).^21,22^

#### Cardiac magnetic resonance (CMR) pre-processing

To test the cross-modal generalizability of the video-based algorithm, we defined a novel pipeline that extracts individual CMR files, identifies long-axis cine views of the left ventricular outflow tract, and compiles these into cine videos. These are then rotated, cropped to cardiac outline, and inverted to grayscale to create clips that loosely mimic the acquisition of a PLAX view on echocardiography (***Supplement***).

#### DASSi calculation

DASSi can be directly quantified using a simple two-dimensional video.^16^ Our algorithm provides a numerical probability of severe AS phenotype ranging from 0 (lowest probability) to 1 (highest probability). For reference, the validated cut-off for *screening of severe* AS in the general population was previously set at 0.607.^16^ Deployment of DASSi involves sequential automated steps that include de-identification, down-sampling, automated view classification, and inference using an ensemble model (***Supplement***). We computed DASSi at the level of each echocardiographic study by averaging video-level predictions, as previously described.^16,23^ For CMR, we applied DASSi directly to the pre-processed PLAX-like CMR videos (one video per patient).

### Definition of outcomes

#### Echocardiographic outcomes

The *primary echocardiographic outcome* was the annualized rate of change in the AV-V_max_ (m/sec/year). If three or more studies were present, we calculated the rate of change as the coefficient of a univariate ordinary least squares regression model of time against AV-V_max_. To avoid outlier effects, we winsorized the rate of change to no less than −1 m/sec/year and no more than +2 m/sec/year.^24^ We chose this over the aortic valve area (AVA) or mean gradient, as these were not consistently measured in patients without AS (missing in 36.8% and 29.0% of cases, respectively). *Secondary echocardiographic outcomes* were: i) progression to a higher severity (mild-moderate-severe) stage, ii) development of any (≥mild) AS (among patients without AS at baseline), iii) and a sensitivity analysis on discrimination of participants with fast (≥0.4 m/sec/year) vs no measured progression (*no increase*) in AV-V_max_.

#### Clinical outcomes

The *primary clinical outcome* was time-to-AVR, with all-cause mortality as a competing risk. These outcomes were derived from the linked institutional electronic health records, which provided dates of relevant procedures and the date of death, including out-of-hospital deaths. Across cohorts, we applied a blanking period of 6 months to avoid contamination of our results from echocardiograms performed during acute illness or from early AVR in the setting of possible AS misclassification. (see ***Supplement***).

### Model explainability

#### Saliency maps

To supplement our prior work in echocardiography,^16^ we generated sample saliency maps for selected high DASSi cases using the self-supervised part of the ensemble model and Gradient-weighted Class Activation Mapping (Grad-CAM).^25^ We present the pixelwise maximum along the temporal axis to capture the most salient regions.

#### Phenome-wide association study (PheWAS)

To explore the clinical correlates of DASSi we applied a computational approach that automatically maps ICD-10 codes to 1,572 unique phenomic concepts, adjusts estimates for relevant covariates (age and sex), applies Bonferroni correction, and visualizes the results using a Manhattan plot.^26,27^

#### Statistical Methods

Categorical variables are summarized as counts (valid percentages), and continuous variables as mean ± standard deviation, or median [25^th^-75^th^ percentile], unless specified otherwise. Normally distributed continuous variables between two groups were compared by unpaired t-test. Pairwise comparisons between continuous variables or an ordinal and a continuous variable were performed using Spearman’s ρ coefficient. Across all multivariable regression models covariates with missing values were imputed using non-parametric chained equation imputation with random forests by including all model covariates (age/sex/race/ethnicity/AV-V_max_/DASSi/baseline LVEF).^28^ Correlations between continuous variables were visualized using Loess regression plots that use local weighted regression to fit smooth curves.

For the analysis of the echocardiographic and clinical outcomes, we defined nested models that included the patient’s age, sex, race, ethnicity, AV-V_max_, and LVEF at baseline, followed by the addition of DASSi. For the rate of change in AV-V_max_, we fit a multivariable generalized linear model (GLM) and compared the improvement in model performance when adding DASSi using the likelihood-ratio (LR) chi-squared test. Calibration was examined based on the alignment between observed progression rates across progressively higher DASSi in the CSMC cohort based on the YNHHS models. The performance of the two models in discriminating individuals with fast vs no progression was examined by comparison of model AUROC by DeLong’s test. Based on the clinical interaction between related AS metrics, we evaluated an interaction term between AV-V_max_, or the dimensionless valve index (DVI), and DASSi and mapped the results using contour plots. We also present tables of sensitivity, specificity, positive (LR+) and negative LR (LR-) for AV-V_max_ progression across DASSi/AV-V_max_ groups.

For the composite outcome of time-to-AVR, time-to-next severity stage, or any AS, we first fit multivariable Cox regression models to account for the variable length of follow-up. The proportionality of hazards was assessed by visualizing Schoenfeld residuals. For the analysis of AVR with death as a competing risk, we fit multivariable Fine-Gray proportional sub-distribution hazard regression models. Using nested models that included baseline predictors with or without DASSi, we assessed the improvement in discrimination and reclassification with DASSi by a LR test, but also C-statistic, categorical net reclassification improvement (NRI) indices (defining low, intermediate and high-risk categories based on a risk of AVR of <5%, 5-20% and ≥20% at the median follow-up) and integrated discrimination improvement (IDI). The latter are only presented in the YNHHS cohort due to limited event counts in the CSMC or UKB cohorts (n=715, versus 56, and 52 events respectively). Calibration curves are presented at t=4 years (rounded median follow-up). We also present adjusted survival curves across DASSi groups, and forest plots for subgroup analyses. The association of the top PheWAS hits with DASSi was explored in age- and sex-adjusted multivariable linear regression models.

All statistical tests were two-sided with a significance level of 0.05 (except for the PheWAS analysis where a Bonferroni correction was applied). Where needed, 95% confidence intervals (CI) were derived from bootstrapping (200 replications). Analyses were performed using Python (version 3.11.2) and R (version 4.2.3).

#### Ethical Approval of Studies and Informed Consent

The study was reviewed by the Yale and Cedars-Sinai Institutional Review Boards (IRBs), which approved the study protocol and waived the need for informed consent as the study represents a secondary analysis of existing data (Yale IRB ID #2000029973). The UK Biobank analysis was conducted under research application #71033.

## RESULTS

### Study overview and population

The YNHHS echocardiography cohort consisted of 8,798 patients (4,250 [48.3%] women) with a median age of 71 [IQR:60-80] years. Patients self-identified with the following race categories: 737 (8.4%) African American, 108 (1.2%) Asian, 6,347 (72.1%) White, and 1,606 (18.3%) other. Moreover, 613 (7.0%) self-identified as Hispanic. At the time of the baseline echocardiographic assessment, 1,047 (11.9%) participants had aortic sclerosis without stenosis, 2,017 (22.9%) mild AS and 979 (11.1%) moderate AS.

The CSMC echocardiography cohort included 3,801 participants (1,685 [44.3%] women) with a median age of 67 [IQR:54-78] years. Patients self-identified with the following race and ethnicity categories: 551 (14.5%) African American, 296 (7.8%) Asian, 424 (11.2%) Hispanic, 2,152 (56.6%) White, and 378 (9.9%) other. At baseline, 3,392 (89.2%) had no AS, 251 (6.6%) sclerosis without stenosis, 83 (2.2%) mild AS, and 75 (2.0%) moderate AS.

From the UK Biobank, we included 45,474 individuals (65 [IQR:59-71] years, 23,559 [51.8%] women), 182 (0.4%) of whom had AS (**Table 1** and **eTable 1**). Patients self-identified with the following race categories: 619 (1.4%) Asian, 304 (0.7%) Black (or Black British), 43,990 (96.7%) White, and 561 (1.2%) other.

**Table 1.**
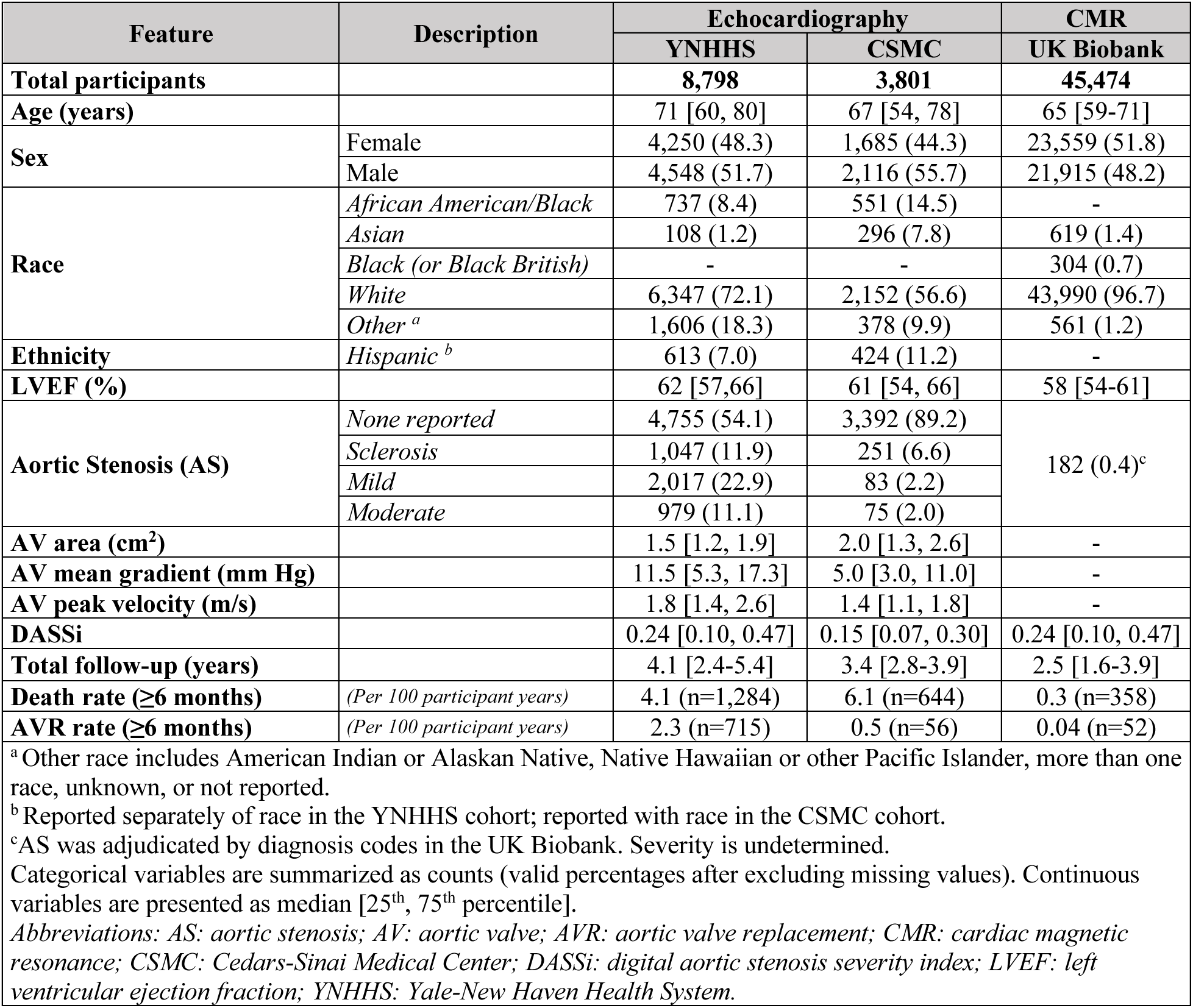
Table of cohort demographics.

### Baseline DASSi phenotyping on echocardiography

In the YNHHS cohort, the median DASSi was 0.24 [IQR:0.10-0.47]. Higher baseline DASSi was associated with greater AS severity by traditional Doppler-derived parameters (AV-V_max_: ρ=0.63, [*n*=8,798]; mean AV gradient: ρ=0.64, [*n*=6,220]; AVA: ρ=-0.53, [*n*=5,410]; and DVI: ρ=-0.63, [*n*=8,163], all *p*<0.001). There was a modest correlation with diastolic dysfunction by E/e’ (ρ=0.36, [*n*=7,079]), left atrial volume index (ρ=0.31, [*n*=7,421]), and right ventricular systolic pressure (ρ=0.18, [*n*=6,312], *p*<0.001 for all three). DASSi was independent of LVEF (ρ=-0.01, [*n*=8,608], *p*=0.39) and stroke volume (ρ=-0.02, [*n*=7,073], *p*=0.11). Collectively, these parameters explained less than half of the variation in DASSi (R^2^=0.46, [95%CI:0.44-0.49]).

### DASSi and echocardiographic progression of AS

#### YNHHS cohort

In total, 5,483 of 8,798 patients (62.3%) in the YNHHS cohort had at least one follow-up study (median 4 [IQR:2-5] studies/patient) within a median of 3.5 [IQR:2.3-4.6] years. DASSi ranged from 0.14 [IQR:0.05-0.27] among patients without AS or sclerosis to 0.61 [IQR:0.48-0.72] among patients with moderate AS (**eFigure 1A-B**).

When stratified by DASSi, the annual change in AV-V_max_ ranged from 0.04±0.01 (standard error of mean) m/sec/year for DASSi values <0.2 at baseline, to 0.21±0.01 for baseline values ≥0.6 (**eFigure 2**), a trend that persisted within each distinct baseline AS stenosis group (**Figure 2A** and **eTable 2**). The addition of DASSi to a model consisting of baseline AV-V_max_, LVEF, age, sex, and race/ethnicity was associated with an +0.033 m/s/year [95%CI:0.028-0.038, *p*<0.001] adjusted annualized AV-V_max_ increase independent of the number of studies and follow-up length (**Figure 2B**) and significantly improved baseline model performance (*p* <0.001 by LR test). There was interaction between baseline DASSi and the flow-corrected DVI (*p*=0.002 for interaction), but not AV-V_max_ (*p*=0.26 for interaction) (**Figure 2C-D**), with higher DASSi associated with faster progression rates for lower baseline DVI. The association between DASSi and AV-V_max_ change remained consistent across demographic subgroups, impaired or preserved left ventricular function (LVEF ≥ vs <50%) and baseline AV-V_max_ strata (**eFigure 3**).

**Figure 2.**
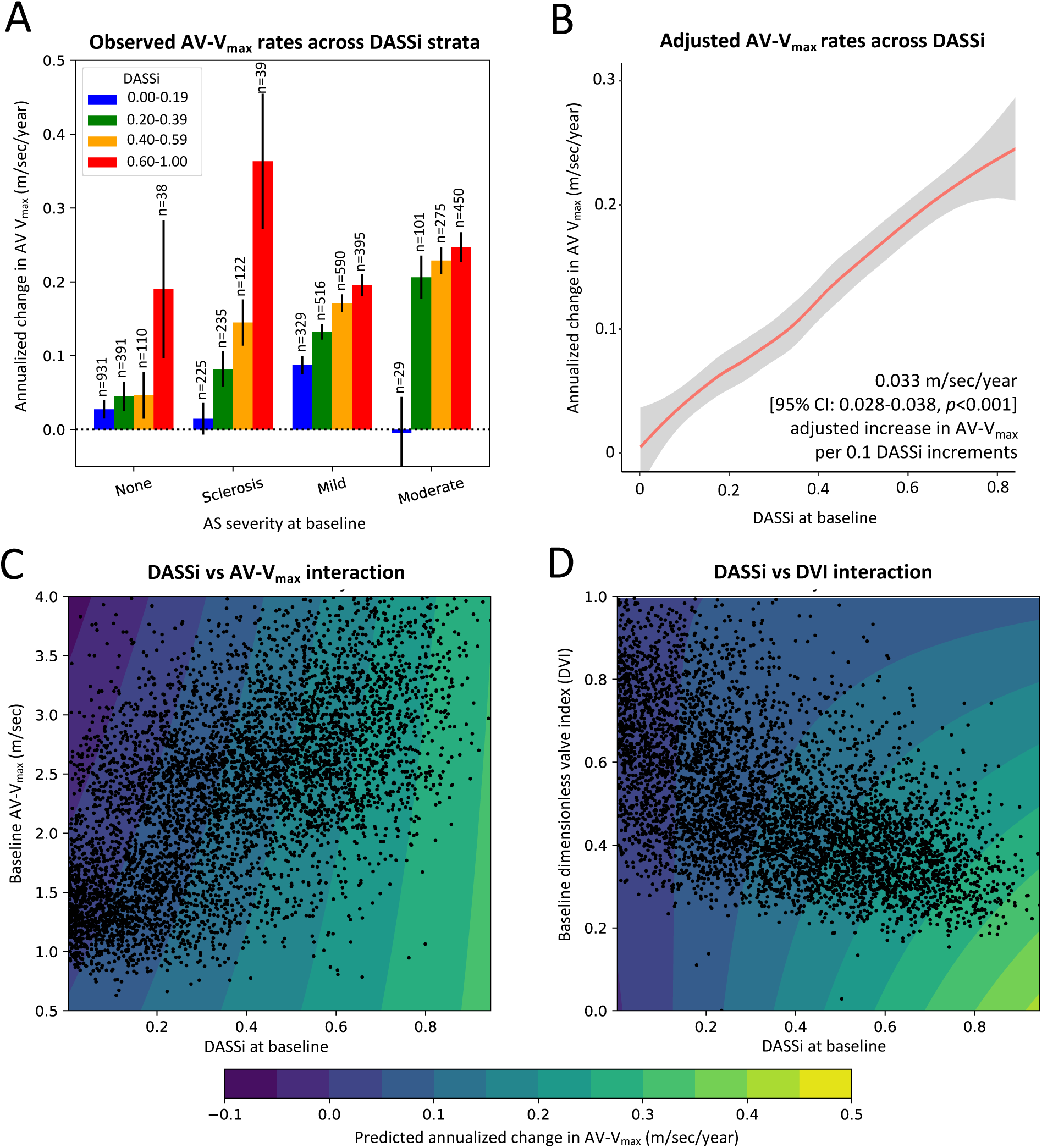
Stratifying the rate of aortic stenosis (AS) progression by DASSi. **(A)** Observed progression rates across DASSi subgroups stratified by baseline severity of AS (error bars denote the standard error of mean) in the YNHHS cohort. The counts of unique observations for each group are provided above each bar. **(B)** Loess regression curves with 95% confidence bands showing the association between baseline DASSi and the annual rate of change in the peak aortic valve velocity (AV-V_max_), adjusted for baseline AV-V_max_, left ventricular ejection fraction, age, sex, ethnicity/race. **(C-D)** Contour plots examining the interaction between DASSi and **(C)** the baseline AV-V_max_, as well as **(D)** the dimensionless valve index (DVI). On these plots, each dot represents a unique observation (participant); the horizontal/vertical axes represent the baseline DASSI/AV-V_max_ values, respectively, and the colored contours denote the adjusted AV-V_max_ progression rates. *AS: aortic stenosis; AV-V_max_: peak aortic valve velocity; DASSi: digital aortic stenosis severity index; YNHHS: Yale-New Haven Health System*.

DASSi improved the baseline model’s ability to discriminate individuals with the fastest progression rates (≥0.4 m/sec/year, *n*=693) from those without progression (*n*=1836), with AUROC increasing from 0.63 to 0.70 (δ[AUROC]=0.07 [95%CI:0.05-0.09]). Among individuals with AV-V_max_<2.5 m/sec, a DASSi of ≥0.5 was linked to LR(+) of 5.9 for identifying participants with AV-V_max_ change ≥0.4 m/sec/year, while for AV-V_max_ 3.0-3.9 m/sec a DASSi <0.2 was associated with LR(-) of 0.1 (**eTable 3**).

In total, 2,037 (37.2%) participants had a follow-up echocardiographic report describing a higher AS severity grade than their baseline study. Greater DASSi was associated with a higher adjusted risk of progressing to a higher severity stage (adj.HR 1.14 [95%CI: 1.12-1.17], *p*<0.001, per 0.1 DASSi increments) after multivariable adjustment including baseline AS severity. DASSi was also positively associated with the future development of *any* AS (*n*=325 new AS cases) in a subset of 2,091 patients without AS at baseline (per 0.1 incr.; adj.HR: 1.16 [95%CI:1.09-1.23], *p*<0.001).

These results were replicated in the geographically distinct CSMC cohort, where 1,292 patients had echocardiographic follow-up over a median of 1.1 [IQR:0.4-1.8] years. DASSi was independently associated with the rate of change in AV-V_max_ (per 0.1 increments: adj. coefficient +0.08 m/s/year [95%CI:0.05-0.11], *p*<0.001; and *p*<0.001 by LR test compared to the baseline model). There was also significant improvement in discrimination of fast vs no progression (n=269 vs 634, respectively; AUROC 0.59 to 0.67; δ[AUROC]=0.08 [95%CI:0.04-0.11]). Higher baseline DASSi was also associated with a higher adjusted risk of progressing to a higher AS stage (*n*=183 cases, adj. HR 1.18 [95%CI:1.08-1.29], *p*<0.001), including progression to new AS (*n*=148 new cases of mild or greater AS among 1208 patients without stenosis at baseline; adj. HR 1.19 [95%CI:1.07-1.33], *p*<0.001), with overall alignment of observed and predicted rates across cohorts and DASSi thresholds (**eFigures 2 and 4**).

### DASSi and future AVR

The 8,798 patients in the YNHHS cohort were followed for 4.1 [IQR:2.4-5.4] years. Following a blanking period of 6 months, 715 underwent AVR. Higher baseline DASSi was independently associated with a higher adjusted risk of AVR (per 0.1 increments; adj.HR 1.21 [95%CI:1.16-1.26], *p*<0.001) (**Table 2, eTable 4,** and **eFigure 5**). Compared with the reference group of DASSi<0.2, the risk of AVR increased from 2.9-fold higher for DASSi 0.2-0.4, to 4.7-fold and 5.4-fold higher for those with DASSi levels 0.4-0.6, and ≥0.6, respectively (**Table 2** and **eFigure 6A**). The prognostic value of DASSi was consistent across sex, age, LVEF, and AV-V_max_ strata (**eFigure 7**), and offered marginal improvements in reclassification and discrimination beyond baseline (*p*<0.001 by LR test; C-statistic 0.896 to 0.902 (δ[C-statistic]: 0.006 [95%CI:0.002-0.009]), categorical NRI 0.05 [95%CI:0.02-0.08], and IDI: 0.006 [95%CI: 0.003-0.009]).

**Table 2.** Adjusted Cox regression estimates for DASSi and late AVR incidence.

| DASSi | Outcome: AVR<sup>a</sup><br>HR [95% CI], *p* value (total counts) |  |  |
| --- | --- | --- | --- |
|  | Echocardiography |  | CMR |
|  | YNHHS cohort<br>(*n*=8,798; 715 events)<sup>b</sup> | CSMC cohort<br>(*n*=3,801; 56 events)<sup>b</sup> | UK Biobank cohort<br>(*n*=45,474; 52 events)<sup>c</sup> |
| Continuous DASSi per 0.1 increments | 1.21 [1.16-1.26], *p*<0.001 | 1.08 [0.92-1.26], *p*=0.36 | 2.09 [1.54-2.83], *p*<0.001 |
| DASSi groups |  |  |  |
| Less than 0.2 | Reference<br>(*n*=3,837) | Reference<br>(*n*=2,302) | Reference<br>(*n*=24,579) |
| 0.2 or greater | 4.97 [2.71-.582], *p*<0.001<br>(*n*=4,961) | 4.04 [0.92-17.70], *p*=0.06<br>(*n*=1,499) | 11.4 [2.56-50.6], *p*=0.001<br>(*n*=20,895) |
| 0.2 to less than 0.4 | 2.86 [1.90-4.30], *p*<0.001<br>(*n*=2,188) | 3.67 [0.79-17.1], *p*=0.10<br>(*n*=881) | 10.7 [2.36-48.8], *p*=0.002<br>(*n*=19,787) |
| 0.4 to less than 0.6 | 4.67 [3.12-6.97], *p*<0.001<br>(*n*=1,601) | 4.68 [1.00-21.87], *p*=0.05<br>(*n*=375) | 24.3[4.64-120.7], *p*<0.001<br>(*n*=1,108) |
| 0.6 or greater | 5.38 [3.54-8.15], *p*<0.001<br>(*n*=1,172) | 3.74 [0.76-18.35], *p*=0.10<br>(*n*=243) | n/a<br>(*n*=0) |
| <sup>a</sup> Estimates for AVR were derived from Fine-Gray proportional sub-distribution hazards regression models with death as competing risk.<br><sup>b</sup> adjusted for baseline age, left ventricular ejection fraction, peak aortic valve velocity, sex, race/ethnicity.<br><sup>c</sup> adjusted for baseline age, left ventricular ejection fraction, aortic stenosis presence, sex, and race/ethnicity.<br>All events were recorded at least 6 months after the index imaging visit (following a 6-month blanking period).<br>AVR: aortic valve replacement; CI: confidence interval; CMR: cardiac magnetic resonance imaging; CSMC: Cedars-Sinai Medical Center;<br>DASSi: digital aortic stenosis severity index; HR: hazard ratio; YNHHS: Yale-New Haven Health System. |  |  |  |

In the CSMC cohort there were 56 AVR procedures performed (>6 months) among 3,801 individuals followed over a median of 3.4 [IQR:2.8-3.9] years. Similar to the YNHHS cohort, when compared with the reference group of DASSi<0.2, those with DASSi ≥0.2 had a 4-fold higher adjusted risk of undergoing AVR at CSMC, though 95% confidence intervals were wide and included the null value (adj.HR 4.04 [95%CI: 0.92-17.70], *p*=0.06) (**eFigure 6B; Table 2**). A YNHHS-based model incorporating age, sex, AV-V_max_, and DASSi showed good calibration in the CSMC cohort with a mean absolute error of 0.6% for AVR at 4 years (**eFigure 8**), whereas the addition of DASSi to the baseline parameters improved the goodness-of-fit of the model (*p*<0.001 by LR test).

#### Cross-modal validation of DASSi using CMR videos

We designed a novel pipeline that enables direct cross-modal application of the original DASSi algorithm to cine CMR videos by creating pseudo-PLAX echo views. Activation maps across two examples with the highest DASSi (0.5 to 0.6) showed that the model focused on myocardial structures, such as the left atrium and right ventricle (**Figure 3A**). CMR-based DASSi successfully discriminated between patients with vs without diagnosed AS (DASSi: 0.32 [IQR:0.25-0.38], *n*=182, vs 0.19 [IQR:0.12-0.26], *n*=45,292, respectively, *p*<0.001) (**Figure 3B**).

**Figure 3.**
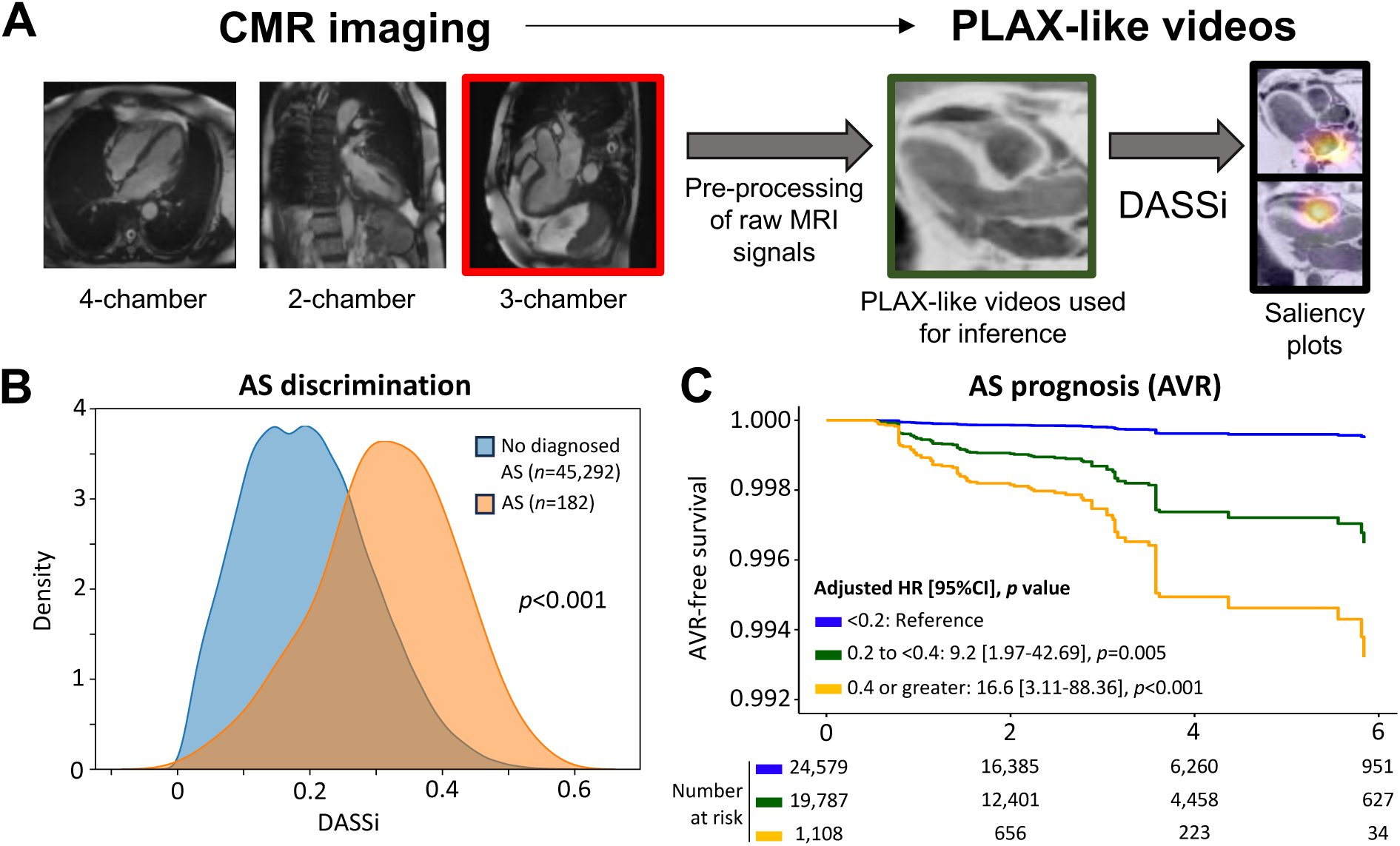
Cross-modal evaluation of DASSi in the UK Biobank. **(A)** We designed a new computational pipeline that enabled the direct transformation of long-axis cine cardiac magnetic resonance (CMR) clips into PLAX-like cine videos. Saliency maps of individuals with the highest DASSi values (0.5-0.59) on CMR demonstrated that the DASSi algorithm focused on key cardiac structures, such as the left atrium and right ventricle. **(B)** Density plot of DASSi (probability that DASSi takes a given value among patients with AS and no AS undergoing CMR imaging in the UK Biobank). **(C)** Flexible adjusted hazard ratio curve for AVR or all-cause mortality based on a multivariable Cox regression model that included DASSi adjusted for age, sex, ethnic background, known AS and baseline left ventricular ejection fraction. *AS: aortic stenosis; AVR: aortic valve replacement; CI: confidence interval; CMR: cardiac magnetic resonance; DASSi: digital aortic stenosis severity index; HR: hazard ratio*.

Over 2.5 [IQR 1.6-3.9] years, there were 52 AVR procedures (>6 months). Compared with the reference group of <0.2, individuals with DASSi levels 0.2-0.4, and 0.4-0.6 had an 11- and 24-fold higher adjusted risk of AVR, independent of age, sex, race/ethnicity, history of AS, and baseline LVEF (**Figure 3C**, **Table 2**). There were no individuals with DASSi values of 0.6 or greater in this cohort.

#### Phenotypic correlates of DASSi

In age- and sex-adjusted PheWAS analyses, DASSi was associated with cardiovascular-specific risk factors – hypertension, atrial fibrillation, obesity, diabetes mellitus, and hypercholesterolemia (**eFigure 9**). Age and sex explained 19% of the variation in DASSi, whereas the addition of the top 10 hits only increased this to 21% (R^2^=0.21). Further adjustment for these phenotypes did not impact the prognostic value of DASSi for AVR (adj.HR 2.12 [95%CI:1.50-2.99], *p*<0.001).

## DISCUSSION

In this multinational cohort study of 58,073 individuals with no or early AS, a novel video-based biomarker of AS – DASSi – was reproducibly associated with the future risk of AS development and progression independent of key clinical parameters and baseline severity by Doppler. The prognostic association of DASSi was shown across three geographically, temporally, and phenotypically distinct cohorts, including patients undergoing clinical echocardiography, the original modality used for its training, as well as a prospective cohort of protocolized CMR imaging in the UK Biobank. These findings support the use of DASSi, a cross-modal AI biomarker for both opportunistic AS screening on echocardiography as well as deeper phenotyping of AS on standard non-invasive modalities and potentially handheld devices.

Efforts to identify individuals at risk of AS progression have been limited by the large burden of milder forms of aortic valve disease, such as aortic sclerosis, found in ~26% of individuals over 65 years.^11,30,31^ However, a challenge in personalizing patient follow-up is the marked variability in the progression rates of patients within similar Doppler-adjudicated severity stages.^32–35^ Traditional risk factors such as hypercholesterolemia, smoking, renal dysfunction, and elevated natriuretic peptides, lack specificity for AS,^15,36^ while alternative Doppler-derived indices require skilled acquisition and modifications to the scanning protocols.^36,37^

Deep learning-enhanced, two-dimensional echocardiography with DASSi aims to bridge this gap by providing a Doppler-independent AS severity metric that can be computed from any portable or standard echocardiogram. Trained to detect generalizable features associated with the severe AS phenotype,^16^ DASSi maintains its prognostic value across the spectrum of AS stages, identifying individuals who do not meet traditional criteria for severe AS, yet exhibit fast rates of progression. Critically, DASSi has several features that make it generalizable and scalable. Unlike prior methods that have utilized structured echo reports and measurements,^37–40^ Doppler images,^41^ or still images of the aortic valve,^42^ DASSi can be directly applied to unprocessed, standard PLAX videos, without the need for any Doppler or two-dimensional measurements, and provides a reader-independent metric to supplement an echocardiographer’s impression, assisting in standardized severity classification.^16^ Furthermore, DASSi can be applied to videos obtained at the point-of-care by individuals with minimal to no training.^16,17^ The value of this approach is supported by consistent and robust effect sizes for both intermediate echocardiographic and long-term clinical outcomes, reproduced across multinational cohorts undergoing both clinically indicated and protocolized imaging.

The cross-modality validation is included not merely to suggest a role for CMR in opportunistic AS screening, but because it shows that DASSi flags a distinct myocardial and valvular phenotypic signature rather than modality- or population-specific confounders.^43^ The specificity of the algorithm is further illustrated by representative saliency maps and PheWAS, suggesting cardiovascular-specific associations with traditional risk factors and links to extra-valvular phenotypes of myocardial remodeling and diastolic dysfunction that may flag AS progression.^44^ However, these only partially explain the variability in DASSi, highlighting the need for multiparametric phenotyping. This innovative framework also suggests a generalizable way to test the validity of new AI-derived echocardiographic biomarkers.

Certain limitations merit consideration. In the echocardiography study, repeat imaging was clinically determined rather than based on a study protocol, thus favoring patients with higher symptom burden and closer interaction with the healthcare system. Reassuringly, our analyses revealed consistent results across varying levels of AS severity at baseline, independent of the frequency of echocardiographic follow-up, for both echocardiographic and clinical outcomes, with the overall findings reproduced in the prospectively enrolled population of the UK Biobank. Further prospective clinical studies will explore the longitudinal association between DASSi and its changes with AS progression across protocol-defined echocardiographic intervals and using multi-modality calibration in racially and ethnically diverse cohorts. Second, it is possible that DASSi flags a cardiovascular phenotype that is not exclusive to AS, a condition that shares several risk factors with coronary artery disease, stroke, and heart failure;^45^ however, analyses across distinct cohorts and outcomes suggest a consistent association with AS-specific outcome measures. Third, outcome ascertainment is the hospital-based echocardiography cohorts was performed through local electronic health records. Thus, events that occurred in different hospitals, or deferral of AVR due to patient preference or ineligibility may not be adequately captured. Indeed, estimates of incremental discrimination and risk stratification were limited by low event counts in two of the three cohorts given the predominance of patience without AS at baseline. Finally, we focused on AV-V_max_, given its more complete capture in our dataset. Though this parameter may be flow-dependent, we demonstrate that associations persisted despite adjusting for LVEF, when modeling against the DVI, and when assessed against future clinical events.

## CONCLUSION

Our study defines a novel AI-based videographic phenotyping of cardiac anatomy and function to detect distinct clinical trajectories among patients with no or non-severe AS, which generalize across multi-national cohorts and non-invasive modalities. By practical deployment in any setting where video-based cardiac imaging is obtained, DASSi may enable more precise risk stratification for the most common valvular disorder without any changes in the image acquisition protocols.

## ARTICLE INFORMATION

### Access to Data and Data Analysis

EKO and RK had full access to all the data in the study and take responsibility for the integrity of the data and the accuracy of the data analysis.

### Data sharing agreement

The UK Biobank structured and imaging data are publicly available for research use (https://www.ukbiobank.ac.uk/enable-your-research/apply-for-access) following an approved access application. The raw clinical echocardiograms from Yale-New Haven Health and Cedars-Sinai Medical Center cannot be made publicly available since they contain personal identifiable information. The code for the analysis can be made available upon reasonable request.

## Data Availability

The echocardiographic videos contain identifiable information and cannot be made available to the public. Analyzed data are available upon reasonable request to the authors.

