## Supplemental Material for "A Multimodality Video-Based AI Biomarker For Aortic Stenosis Development And Progression"

### Table of contents

Supplemental Methods 2
eTable 1 | Demographics and echocardiographic characteristics in the YNHHS and CSMC cohorts by AS stage. 6
eTable 2 | Percentage of participants with progression in AV-$V_{max}$ stratified by DASSi. 7
eTable 3 | Positive and negative likelihood ratios across baseline DASSi thresholds and AV-$V_{max}$ groups for discrimination of rapid AV-$V_{max}$ progression (≥ 0.4 vs <0.4 m/sec/year). 8
eTable 4 | Association between DASSi and AV-$V_{max}$ with AVR. 9
eFigure 1 | Baseline DASSi phenotyping and observed rates of AS progression by AV-Vmax in the YNHHS cohort. 10
eFigure 2 | Echocardiographic progression of aortic stenosis across baseline DASSi groups. 11
eFigure 3 | Subgroup analysis of the association between baseline DASSi and echocardiographic aortic stenosis (AS) progression in the YNHHS cohort. 12
eFigure 4 | Predicted versus observed rates of AV-Vmax progression across baseline DASSi levels. 13
eFigure 5 | Plots of scaled Schoenfeld residuals. 14
eFigure 6 | Association of baseline echocardiographic DASSi phenotypes with future AVR. 15
eFigure 7 | Subgroup associations between baseline echocardiographic DASSi phenotypes and future AVR in the YNHHS cohort. 16
eFigure 8 | Internal and external calibration curves for a DASSi-based model to predict future AVR. 17
eFigure 9 | Phenome-wide association of CMR-derived DASSi. 18
Supplemental References 19

**Supplemental Methods**

### 1. Cohort Definition

***Yale-New Haven Health System (YNHHS) cohort***: We identified individuals who had undergone a baseline echocardiogram from 2015 to 2021 at one of five YNHHS-affiliated hospitals (Yale-New Haven, Bridgeport, Lawrence & Memorial and Greenwich Hospitals in Connecticut, USA, & Westerly Hospital in Rhode Island, USA) or one of the affiliated outpatient sites. The original YNHHS cohort included all (n=6,931) studies from the original deep learning model report, where mild and moderate AS had previously been oversampled by 5-fold. We limited our analysis to studies used in the validation, internal and external testing samples.[1] This was done to avoid data leakage from the model's original training set. We further enriched the study sample with studies (n=4,447) of patients without severe AS at baseline from the same study period. We included individuals: i) with baseline peak aortic valve velocity (AV Vmax) of less than 4 m/sec at baseline, ii) without history of AVR at baseline, and iii) with studies that included PLAX videos. After applying these criteria, 8,798 unique patients were included in the analysis, for whom we retrieved all follow-up echocardiographic reports, procedures, and death reports.

***Cedars-Sinai Medical Center (CSMC) cohort***: For further testing in an additional geographically distinct cohort, all transthoracic echocardiograms performed at CSMC (Los Angeles, California, USA) between 1 January 2018 and 31 December 2019 were retrieved, as previously reported.[1] After excluding studies with prosthetic aortic valves, 4,000 TTEs were sampled at random. After applying the same criteria as the YNHHS cohort, 3,801 individuals with baseline TTE were included in the study.

***UK Biobank (UKB) cohort***: The UK Biobank is a prospective observational study of 502,468 participants aged 40-69 years who were recruited between 2006 and 2010 which continues to collect extensive phenotypic and genotypic details using multimodal data capture.[2] Though the UK Biobank is not perfectly representative of the broader UK population,[3] its size, accuracy, depth of phenotypic and genomic characterization, and prospective nature have identified it as a valuable source for epidemiological research and validation of risk stratification tools in overall healthy community-dwelling individuals, that are often under-represented in hospital-based cohorts. We focused our analysis on 45,479 individuals who were enrolled in the imaging assessment by CMR,[4] which was performed between 2014 and 2020. We included eligible individuals who underwent cardiac MRI imaging as part of a follow-up comprehensive imaging visit, using a detailed protocol that has been previously described.[4] This allowed us to expand our analysis to individuals who underwent imaging independent of symptoms, thus minimizing confounding by indication, while alleviating potential confounding effects from signals that overfit to technical aspects of echocardiography. After excluding individuals who had withdrawn consent, those with AVR prior to CMR imaging and those for whom the files could not be processed to generate videos, we included 45,474 eligible individuals.

### 2. Interpretation Of Clinical Echocardiograms

**AS severity:** In both the YNHHS and CSMC cohorts, the presence of AS severity was adjudicated based on the original echocardiographic report and reflected the final severity grade assigned by the interpreting physician. This was adjudicated by the reading physician based on an integrated assessment of traditional echocardiographic and Doppler criteria, such as peak aortic valve (stenosis) jet velocity, mean transaortic/trans-valvular gradient, and mean valve area, as assessed by the continuity equation. According to the guidelines, cut-offs of ≥4 m/sec, ≥40 mm Hg and less than <1.0 $cm^2$, respectively, were consistent with severe AS.[5] However the final determination regarding the presence and severity of AS was made by the interpreting physician who had access to all images and measurements and signed the final clinical report. It should also be noted that even though Doppler-derived parameters may have been missing from the structured database, the interpreting physician had access to all required images and Doppler recordings to make a determination about the presence and/or severity of AS. Cases classified as "mild-moderate", or "moderate-severe", were grouped with the "mild" and "moderate" severity groups, respectively, as per our previous development study where they were included in the control set.[1] Cases with low-flow, low-gradient severe AS or paradoxical AS, were included in the "severe" group, consistent with our prior work, and were therefore excluded from our subsequent analysis.[1]

***LVEF (left ventricular ejection fraction):*** The LVEF label was obtained from the final echocardiographic report and was assigned by the interpreting physician using one of the following methods: i) three-dimensional (3D) echocardiography, ii) 2D echocardiography based on the Simpson's biplane method; or iii) a range by visual assessment, reported in 5% increments (i.e., <5%, 5-10%, 10-15%, 20-25%, 25-30%, … 70% or greater). If the LVEF was reported as a range, then we used the median of the range (e.g. 52.5% for a range of 50-55%).

***Doppler-derived parameters and the peak velocity ratio (dimensionless valve index)*****:** Doppler-derived parameters were extracted from the final echocardiographic report. To account for between-study variability in the Doppler angle or flow states, we also calculated the peak velocity ratio (also known as dimensionless valve index), defined as the ratio of the peak velocity in the left ventricular outflow tract (LVOT Vmax) to the AV $V_{max}$. This was chosen over the ratio of the velocity time integrals (VTI) to minimize data missingness.

### 3. Pre-processing Of Echocardiographic Videos

Deployment of the DASSi model in echocardiographic studies involves the input of a full study, which is de-identified, down-sampled, and then processed for automated view classification to identify the specific videos from each study that correspond to PLAX views. The down-sampled 16-frame clips extracted from 2D PLAX videos are processed in a 3D-ResNet18 network architecture trained to detect severe AS, and predictions are based on an ensemble of three models with a combination of three initializations: random, Kinetics-400, and self-supervised learning for echocardiograms.[6] Model-specific study-level predictions represent the average predictions across all PLAX videos in a study for a given model. Finally, study-level predictions are averaged to form an ensemble, with the output (DASSi) reflecting the probability (from 0 to 1) of a severe AS phenotype across all videos of a given study. Further information on the method development as well as the use of self-supervised learning for echocardiographic model training have been previously reported.[1,6]

### 4. Cardiac Magnetic Resonance (CMR) Study Interpretation

In the UK Biobank, participants underwent a 20-minute cardiac magnetic resonance (CMR) protocol without a pharmacological stressor or contrast agent, which was integrated into a 30-minute combined CMR and abdominal magnetic resonance imaging (MRI) protocol performed using a clinical wide bore 1.5 Tesla scanner (MAGNETOM Aera, Syngo Platform VD13A, Siemens Healthcare, Erlangen, Germany).[4] The UK Biobank CMR protocol was performed as part of a protocolized research protocol and there was no clinical reporting. The left ventricular ejection fraction measurements were derived from the structured dataset that has been made publicly available. We took the average of the following data-fields, or the absolute value of one if the other one was missing: data-fields 22420: "LV ejection fraction",[7] and 24103: "LV ejection fraction",[8] derived through automated measurements without strict quality control as reported in the respective references.

### 5. Pre-Processing Of CMR Cine Videos

Imaging biomarkers are often developed and confined within a single modality, however those that are built on the detection of pertinent anatomical and functional features should exhibit modality-invariance. Proving the prognostic value of a biomarker across two or more modalities is important, since i) it supports the notion that the algorithm learns key representations of the disease, rather than technical confounders specific to the acquisition process; ii) it maximizes the value of AI algorithms by enabling zero-shot predictions in new cohorts and clinical settings; iii) it augments our ability to opportunistically risk stratify for AS using existing data streams. We tested this hypothesis by translating DASSi to CMR imaging using the UK Biobank registry.

We restricted our analysis to long axis cines for cardiac function through the left ventricular outflow tract view, which includes the same anatomical structures as the echocardiographic PLAX view which was used for the development of DASSi on echocardiography.[4] Next, we defined an automatic pipeline that extracts individual .DICOM files from each study-specific folder, identifies long axis cine views of the LVOT tract using the available .DICOM headers that suggest a 3-chamber view (CINE-segmented-LAX-3Ch), windows the grayscale according to the default center and width of each study, converts the windowed data to 8-bit .avi files while selecting every other frame (thus creating 25-frame-long clips, by skipping every other clip in the original 50 frames of each view-specific cine) and down-samples to 112 x 112 pixels. The sagittal clips are then rotated by 90 degrees to match the orientation of a PLAX view, cropped (removing 30 pixels from the new left/right side, 20 pixels from the top and 30 pixels from the bottom) to remove structures traditionally not seen on echocardiography, and the grayscale is then inversed to ensure that the myocardial wall appears brighter than its cavity. This enables an MRI2echo transition that does not modify the composition of the underlying signal. The final clips, resampled to 112 x 112 pixels, were then used for direct DASSi inference.

### 6. Key definitions

***Aortic valve replacement (AVR) or valvuloplasty procedures:*** In the YNHHS echocardiography cohort, this was defined based on ICD-9 or ICD-10 codes. We included all ICD-10 codes starting with "02RF", which included all prosthetic valve types via transcatheter or open approach. We also included ICD-10 codes starting with "”027F" to account for valvuloplasty/dilation procedures. For ICD-9 codes, we included codes: "35.01", "35.11", "35.21", "35.21", "35.22", "35.96". In the CSMC cohort AVR procedures were identified through linkage with the local electronic health record and thoracic surgery databases. Surgical AVR cases performed for severe aortic regurgitation or aortic valve endocarditis without prior evidence of AS were censored at the time of surgery and were not included as cases in the outcome analysis. In the UK Biobank (OPCS3/4 codes), we used the following codes: "K26", "K261", "K262", "K263", "K264", "K265", "K268", "K269", "K302", "K312", "K322", "K352", "313.2".

***Aortic stenosis (AS) history:*** In the YNHHS and CSMC cohorts, this was defined based on the echocardiographic report of the corresponding echocardiographic study. In the UK Biobank, and in the absence of aortic stenosis-specific measurements, this was adjudicated based on the presence of an ICD-10 code reflective of aortic stenosis: 'I350', 'I352', 'I060', 'I062'. Since the CMR study was not protocolled for AS and no clinical reporting was performed, we were unable to formally adjudicate the presence/severity of AS in these patients based on cross-sectional imaging data.

***Clinical outcomes:*** For the YNHHS cohort, outcomes were assessed until April 18, 2023. AVR was defined based on procedure codes corresponding to percutaneous or open AVR with any valve type or valvuloplasty/aortic valve dilation. Both in-hospital and out-of-hospital death reports were obtained from the vital statistics log maintained by the health system, drawn from social security administration and state vital statistic records. In the CSMC cohort, mortality data were available until January 1, 2023, transcatheter-only AVR data until May 24, 2022, and surgical AVR data until December 31, 2021. In the UK Biobank, deaths were adjudicated through linkage to national death registries (data-field 40000; "Date of Death"), whereas AVR was defined using operative procedures (data-field 41273; "Operative procedures – OPCS3" and data-field 41272; "Operative procedures – OPCS4") and the associated dates. The date of the last recorded death/procedure in the available data ("2020-12-03") was used for right censoring of the remaining observations.

### 7. Saliency maps

We used the Grad-CAM (Gradient-weighted Class Activation Mapping) method,[23] as previously described in our recent work,[1] to produce saliency maps that provide a visual interpretation of the deep learning model's outputs. Briefly, saliency maps emphasize the areas of the input video that are most influential when a neural network provides inference on a given label (i.e., aortic stenosis phenotype in this manuscript), as determined by the gradient/weight of each input pixel. Here, heatmaps were created for each frame of an echocardiographic video scaled to the resolution that is required by the deep learning model (112 x 112 pixels). We then took the maximal value for each pixel across the time dimension, thus providing a graphical summary of which sections of the image contribute the most to the model's inference.

**eTable 1 | Demographics and echocardiographic characteristics in the YNHHS and CSMC cohorts by AS stage.**

| | YNHHS cohort | | | | | CSMC cohort | | | | |
|---|---|---|---|---|---|---|---|---|---|---|
| | *Missing* | *No AS reported* | *Sclerosis without Stenosis* | *Mild AS* | *Moderate AS* | *Missing* | *No AS reported* | *Sclerosis without Stenosis* | *Mild AS* | *Moderate AS* |
| **Total counts (n)** | | 4755 | 1047 | 2017 | 979 | | 3392 | 251 | 83 | 75 |
| **Age** (years) | - | 65 [55,76] | 75[67,82] | 75 [68,82] | 77 [69,84] | - | 65 [52,76] | 75 [68,82] | 80 [73.50,87] | 84 [75.50,89] |
| **Sex (female)** | - | 2411 (50.7) | 515 (49.2) | 916 (45.4) | 408 (41.7) | - | 1500 (44.7) | 111 (44.2) | 44 (53.0) | 30 (40) |
| **African American race** | 1091 | 519 (12.5) | 70 (7.5) | 105 (6.0) | 43 (5.0) | 106 | 507 (15.4) | 32 (13.0) | 8 (9.8) | 4 (5.7) |
| **Asian race** | | 77 (1.9) | 10 (1.1) | 15 (0.9) | 6 (0.7) | | 266 (8.1) | 22 (8.9) | 3 (3.7) | 5 (6.9) |
| **White race** | | 3201 (77.1) | 794 (85.4) | 1571 (89.5) | 781 (90.0) | | 1890 (57.4) | 151 (61.1) | 56 (68.3) | 55 (75.3) |
| **Other race**[a] | | 356 (8.6) | 56 (6.0) | 65 (3.7) | 38 (4.4) | | 245 | 16 | 6 | 5 |
| **Hispanic ethnicity**[b] | 1334 | 415 (10.3) | 59 (6.5) | 98 (5.8) | 41 (5.0) | | 385 (11.7) | 26 (10.5) | 9 (11.0) | 4 (5.5) |
| **LVIDd Index** ($cm/m^2$) | 1642 | 2.4 [2.2,2.6] | 2.4 [2.2,2.6] | 2.4 [2.2,2.6] | 2.3 [2.1,2.6] | | 2.4 [2.1,2.7] | 2.3 [2.1,2.7] | 2.5 [2.2,2.9] | 2.4 [2.2,2.7] |
| **LA Vol. Index** ($cm^3/m^2$) | 1377 | 29 [22,38] | 33 [25,44] | 34 [27,43] | 35 [27,46] | 616 | - | - | - | - |
| **LA Area Index** ($cm^2/m^2$) | - | - | - | - | - | 808 | 10.2 [8.4,12.7] | 10.8 [8.6,13.6] | 12.4 [9.9,16.7] | 11.9 [9.4,15.1] |
| **RVSP (mmHg)** | 2486 | 28 [22,35] | 30 [24,39] | 30 [25,37] | 30 [26,38] | 2405 | 27 [21,37] | 33 [25,43] | 29 [24,39] | 35 [28,47] |
| **E/E' Avg** | 1719 | 9 [7,12] | 11 [9,15] | 12 [9,16] | 13[10,17] | - | - | - | - | - |
| **LVEF (%)** | 190 | 62 [56,66] | 62 [56,66] | 63 [58,67] | 63 [58,68] | 0 | 61 [55,66] | 60 [52,67] | 61 [54,66] | 62 [50,66] |
| **AV area** (cm2) | 3388 | 1.9 [1.4,2.6] | 1.6 [1.4,2.0] | 1.5 [1.2,1.7] | 1.0 [0.9,1.2] | 2510 | 2.1 [1.3,2.6] | 2.1 [1.7,2.6] | 1.5 [1.4,1.8] | 1.1 [0.9,1.3] |
| **AV Mean Grad.** (mmHg) | 2578 | 5 [4,11] | 7 [5,10] | 14 [11,16] | 24 [21,29] | 2146 | 5 [3,9] | 5 [4,8] | 11 [8,14] | 20 [13,24] |
| **AV Peak Velocity** (m/sec) | - | 1.4 [1.2,1.8] | 1.7 [1.4,2.04] | 2.5 [2.3,2.8] | 3.3 [3.1,3.6] | 914 | 1.3 [1.1,1.7] | 1.4 [1.2,1.7] | 2.3 [2.0,2.5] | 2.8 [2.4,3.2] |
| **DVI** (by peak velocity) | 635 | 0.69 [0.56,0.80] | 0.59 [0.48,0.70] | 0.43 [0.37,0.50] | 0.31 [0.27,0.36] | 980 | 0.73 [0.58,0.84] | 0.71 [0.59,0.81] | 0.47 [0.40,0.55] | 0.34 [0.28,0.40] |
| **DASSi** | - | 0.14 [0.05,0.27] | 0.24 [0.11,0.39] | 0.42 [0.25,0.58] | 0.61 [0.48,0.72] | 0 | 0.14 [0.06,0.27] | 0.19 [0.11,0.31] | 0.44 [0.29,0.56] | 0.57 [0.47,0.70] |
| **Deaths (≥6 months)** | - | 382 | 120 | 286 | 168 | - | 281 | 21 | 14 | 15 |
| **AVR events (≥6 months)** | - | 66 | 11 | 246 | 392 | - | 42 | 0 | 11 | 3 |

[a] Other race includes American Indian or Alaskan Native, Native Hawaiian or other Pacific Islander, more than one race, or not reported.

[b] Reported separately of race in the YNHHS cohort; reported with race in the CSMC cohort.

Categorical variables are summarized as counts (valid percentages). Continuous variables are presented as median [$25^{th}$, $75^{th}$ percentile]. *AS: aortic stenosis; AV: aortic valve; CSMC: Cedars-Sinai Medical Center; DASSi: digital aortic stenosis severity index; E/e': early diastolic transmitral flow velocity to tissue Doppler mitral anular early diastolic velocity; LA: left atrium; LVEF: left ventricular ejection fraction; LVIDd: left ventricular internal diastolic dimeter; MRI: magnetic resonance imaging; RVSP: right ventricular systolic pressure; YNHHS: Yale-New Haven Health System.*

**eTable 2 | Percentage of participants with progression in AV-$V_{max}$ stratified by DASSi.**

| | YNHHS cohort | | | | CSMC Cohort | | | |
|---|---|---|---|---|---|---|---|---|
| **Threshold for AV $V_{max}$ change** | **DASSi <0.2** | **DASSi 0.2-0.39** | **DASSi 0.4-0.59** | **DASSi ≥0.6** | **DASSi <0.2** | **DASSi 0.2-0.39** | **DASSi 0.4-0.59** | **DASSi ≥0.6** |
| **>0 m/sec/year** | 913 (54.5%) | 950 (64.5%) | 980 (74.3%) | 794 (78.2%) | 367 (49.2%) | 136 (48.7%) | 92 (57.5%) | 63 (58.9%) |
| **>0.1 m/sec/year** | 527 (31.4%) | 607 (41.2%) | 711 (53.9%) | 607 (59.8%) | 251 (33.6%) | 105 (37.6%) | 77 (48.1%) | 55 (51.4%) |
| **>0.2 m/sec/year** | 316 (18.9%) | 364 (24.7%) | 460 (34.9%) | 437 (43.1%) | 177 (23.7%) | 77 (27.6%) | 64 (40.0%) | 45 (42.1%) |
| **>0.3 m/sec/year** | 195 (11.6%) | 224 (15.2%) | 304 (23.0%) | 316 (31.1%) | 149 (20.0%) | 67 (24.0%) | 55 (34.4%) | 41 (38.3%) |
| **>0.4 m/sec/year** | 123 (7.3%) | 148 (10.0%) | 200 (15.2%) | 222 (21.9%) | 121 (16.2%) | 59 (21.1%) | 53 (33.1%) | 36 (33.6%) |

*AS: aortic stenosis; AV-$V_{max}$: peak aortic valve velocity; CSMC: Cedars-Sinai Medical Center; DASSi: digital aortic stenosis severity index; YNHHS: Yale-New Haven Health System.*

**eTable 3 | Positive and negative likelihood ratios across baseline DASSi thresholds and AV-$V_{max}$ groups for discrimination of rapid AV-$V_{max}$ progression (≥ 0.4 vs <0.4 m/sec/year).**

| DASSi threshold | Participants below threshold (n, %) | Sensitivity (95% CI) | Specificity (95% CI) | Positive LR(+) (95% CI) | Negative LR(-) (95% CI) |
|---|---|---|---|---|---|
| **Baseline AV Vmax <2.5 m/sec** | | | | | |
| **0.2** | 781 (53.6) | 0.62 (0.57-0.65) | 0.58 (0.56-0.61) | 1.47 (1.36-1.57) | 0.66 (0.6-0.74) |
| **0.3** | 1127 (77.4) | 0.41 (0.35-0.47) | 0.82 (0.8-0.84) | 2.34 (1.92-2.67) | 0.71 (0.64-0.79) |
| **0.4** | 1322 (90.7) | 0.2 (0.16-0.25) | 0.94 (0.93-0.95) | 3.24 (2.56-4.48) | 0.85 (0.79-0.89) |
| **0.5** | 1426 (97.9) | 0.06 (0.04-0.08) | 0.99 (0.98-0.99) | 5.86 (3.4-11.8) | 0.95 (0.94-0.97) |
| **Baseline AV Vmax 2.5-2.9 m/sec** | | | | | |
| **0.2** | 127 (22.9) | 0.92 (0.88-0.96) | 0.3 (0.26-0.35) | 1.31 (1.23-1.48) | 0.27 (0.12-0.43) |
| **0.3** | 254 (45.9) | 0.73 (0.67-0.77) | 0.55 (0.51-0.59) | 1.62 (1.4-1.9) | 0.49 (0.41-0.63) |
| **0.4** | 408 (73.7) | 0.38 (0.32-0.42) | 0.79 (0.75-0.83) | 1.77 (1.41-2.12) | 0.79 (0.75-0.87) |
| **0.5** | 518 (93.5) | 0.11 (0.06-0.15) | 0.96 (0.93-0.98) | 2.4 (1.23-5.12) | 0.93 (0.88-0.99) |
| **Baseline AV Vmax 3.0 -3.9 m/sec** | | | | | |
| **0.2** | 47 (8.9) | 0.99 (0.97-1.0) | 0.14 (0.11-0.18) | 1.14 (1.1-1.2) | 0.11 (0.03-0.22) |
| **0.3** | 132 (25.0) | 0.89 (0.86-0.92) | 0.34 (0.29-0.38) | 1.35 (1.26-1.47) | 0.33 (0.23-0.42) |
| **0.4** | 257 (48.7) | 0.67 (0.62-0.74) | 0.59 (0.55-0.64) | 1.63 (1.44-1.9) | 0.56 (0.44-0.67) |
| **0.5** | 416 (78.8) | 0.29 (0.24-0.36) | 0.84 (0.82-0.89) | 1.85 (1.49-2.72) | 0.84 (0.75-0.9) |

*AV-$V_{max}$: peak aortic valve velocity; CI: confidence interval; DASSi: digital aortic stenosis severity index; LR: likelihood ratio.*

**eTable 4 | Association between DASSi and AV-$V_{max}$ with AVR.**

| Independent variable | AVR[a] (HR [95% CI], *p* value) | | |
|---|---|---|---|
| | **AV-$V_{max}$ <2.5 m/sec** | **AV-$V_{max}$ 2.5-2.9 m/sec** | **AV-$V_{max}$ 3.0-3.9 m/sec** |
| **DASSi (per 1SD incr.)** | 2.23 [1.36-3.68], *p*=0.002 | 1.30 [1.13-1.50], *p*<0.001 | 1.39 [1.25-1.55], *p*<0.001 |
| **AV-$V_{max}$ (per 1 SD incr.)** | 3.71 [1.62-8.54], *p*=0.002 | 1.33 [1.15-1.53], *p*=0.04 | 1.21 [1.11-1.33], *p*<0.001 |

[a] Estimates were derived from Fine-Gray proportional subdistribution hazards regression models accounting for the competing risk of death. All events were recorded following a 6-month blanking period in the YNHHS cohort. *AV-$V_{max}$: peak aortic valve velocity; AVR: aortic valve replacement; CI: confidence interval; DASSi: digital aortic stenosis severity index; HR: hazard ratio; SD: standard deviation; YNHHS: Yale-New Haven Health System.*

### eFigure 1

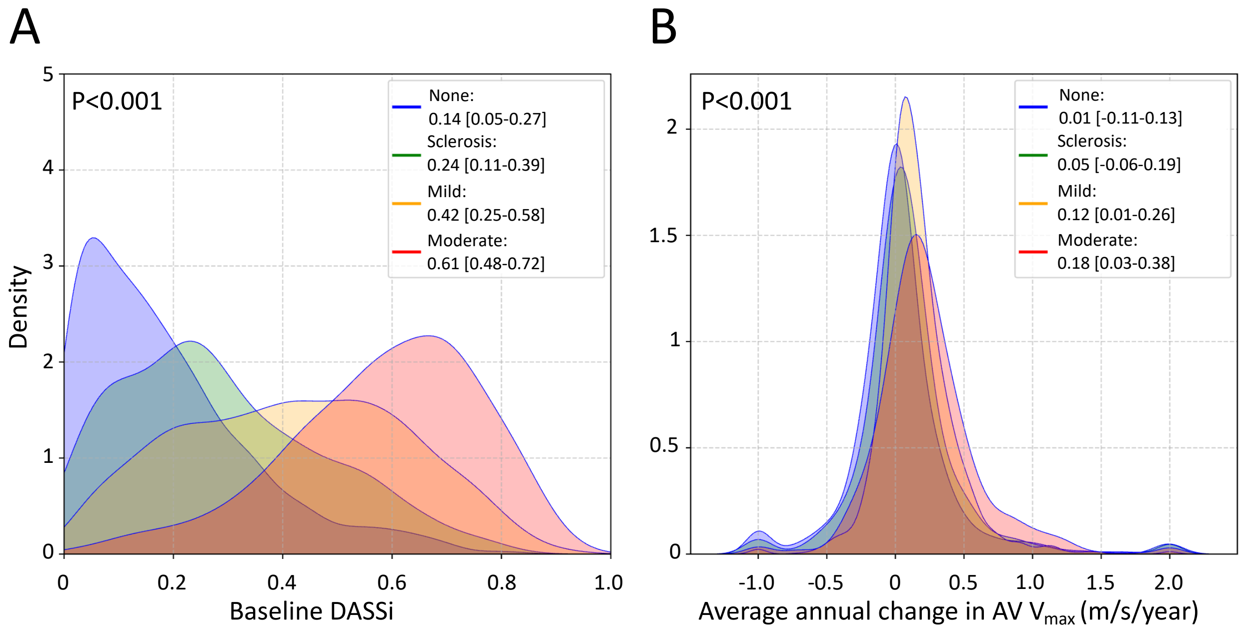


**eFigure 1 | Baseline DASSi phenotyping and observed rates of AS progression by AV-Vmax in the YNHHS cohort. (A)** Density plot of the DASSi (digital aortic stenosis [AS] severity index) at baseline across AS severity groups. **(B)** Density plots of the observed, annualized rate of change in the peak aortic valve velocity across time (in m/sec/year) stratified by the baseline AS severity group. *AV-$V_{max}$: peak aortic valve velocity; AS: aortic stenosis; DASSi: digital aortic stenosis severity index; YNHHS: Yale-New Haven Health System.*

### eFigure 2

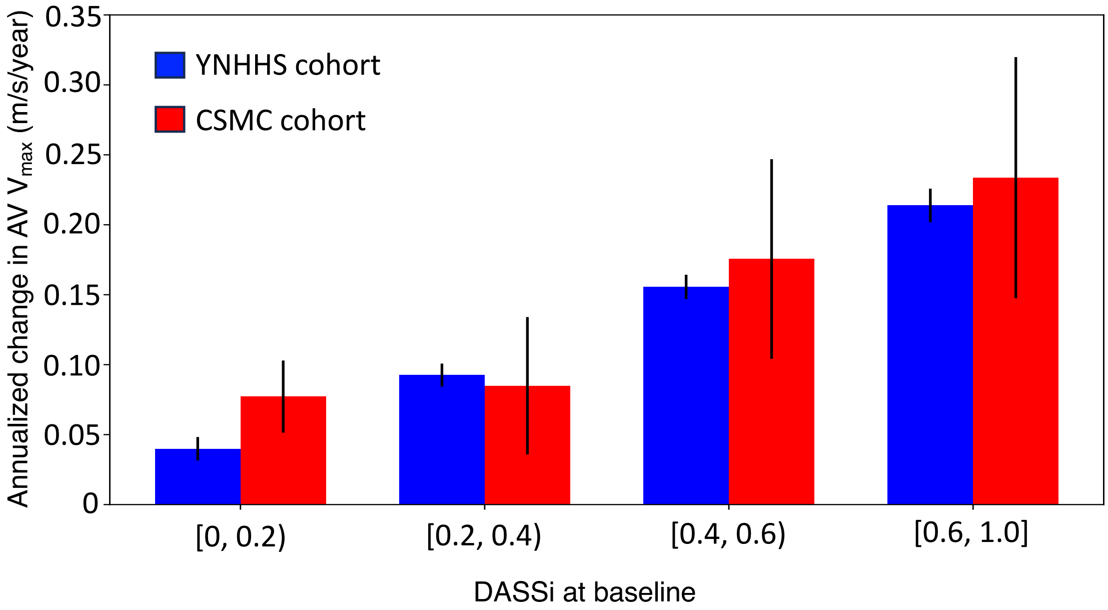


**eFigure 2 | Echocardiographic progression of aortic stenosis across baseline DASSi groups.** Unadjusted rates of progression in AV-$V_{max}$ (peak aortic valve velocity) across baseline DASSi subgroups in the YNHHS and CSMC cohorts. Bars denote mean rates with corresponding error bars reflecting the standard errors of mean. *AV-Vmax: peak aortic valve velocity; CSMC: Cedars-Sinai Medical Center; DASSi: digital aortic stenosis severity index; YNHHS: Yale-New Haven Health system.*

**eFigure 3**

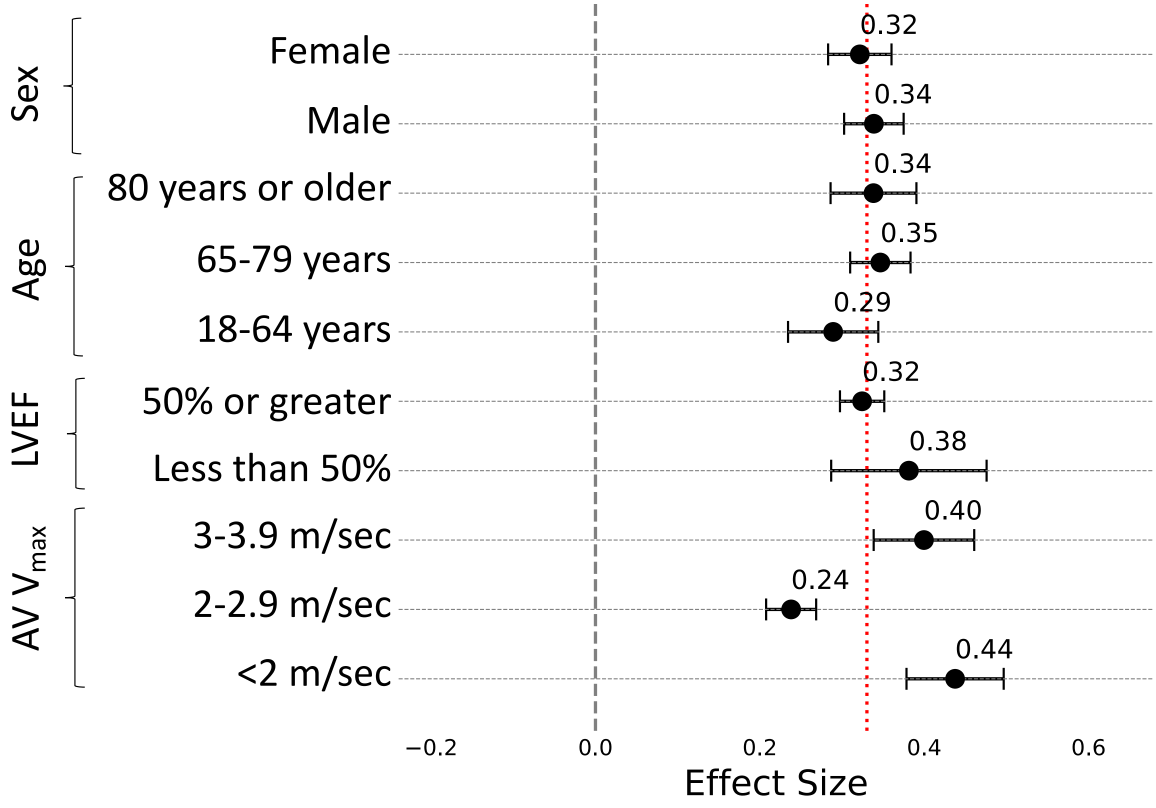


**eFigure 3 | Subgroup analysis of the association between baseline DASSi and echocardiographic aortic stenosis (AS) progression in the YNHHS cohort.** Forest plot describing the association between baseline DASSi (0 through 1) and the annualized rate of change in the peak aortic valve velocity (AV-$V_{max}$), across key subgroups. The graph illustrates the regression coefficients with the corresponding 95% confidence interval. *AV-$V_{max}$: peak aortic valve velocity; DASSi: digital aortic stenosis severity index; EF: ejection fraction; YNHHS: Yale-New Haven Health System.*

### eFigure 4

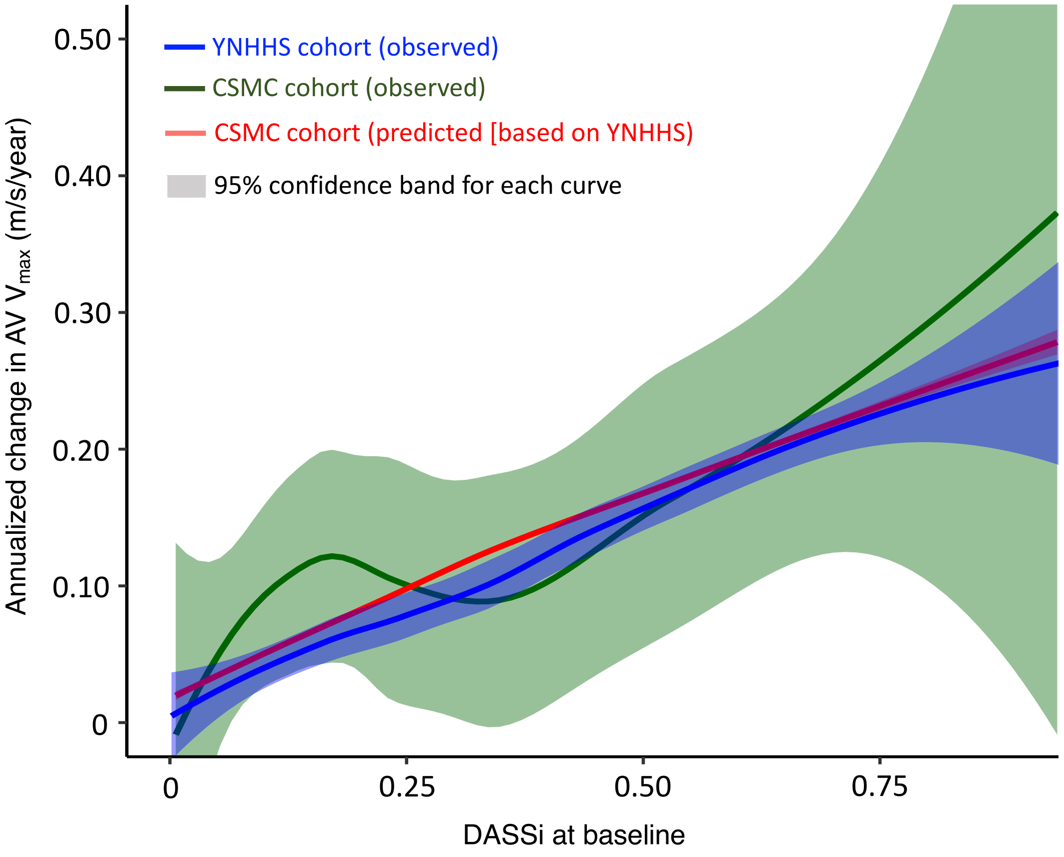


**eFigure 4 | Predicted versus observed rates of AV-Vmax progression across baseline DASSi levels.** Loess regression plot demonstrating the association between baseline DASSi and the rate for echocardiographic progression in the AV-$V_{max}$. Loess regression utilizes nonparametric modeling that enables local weighted regression to fit a smooth curve that can illustrate local patterns in data. The curve demonstrates similar trends across both cohorts (YNHHS and CSMC; blue and green, respectively) with overlapping 95% confidence bands for both observed and predicted values (in CSMC; green and red, respectively). *AV-$V_{max}$: peak aortic valve velocity; CSMC: Cedars-Sinai Medical Center; DASSi: digital aortic stenosis severity index; YNHHS: Yale-New Haven Health System.*

**eFigure 5**

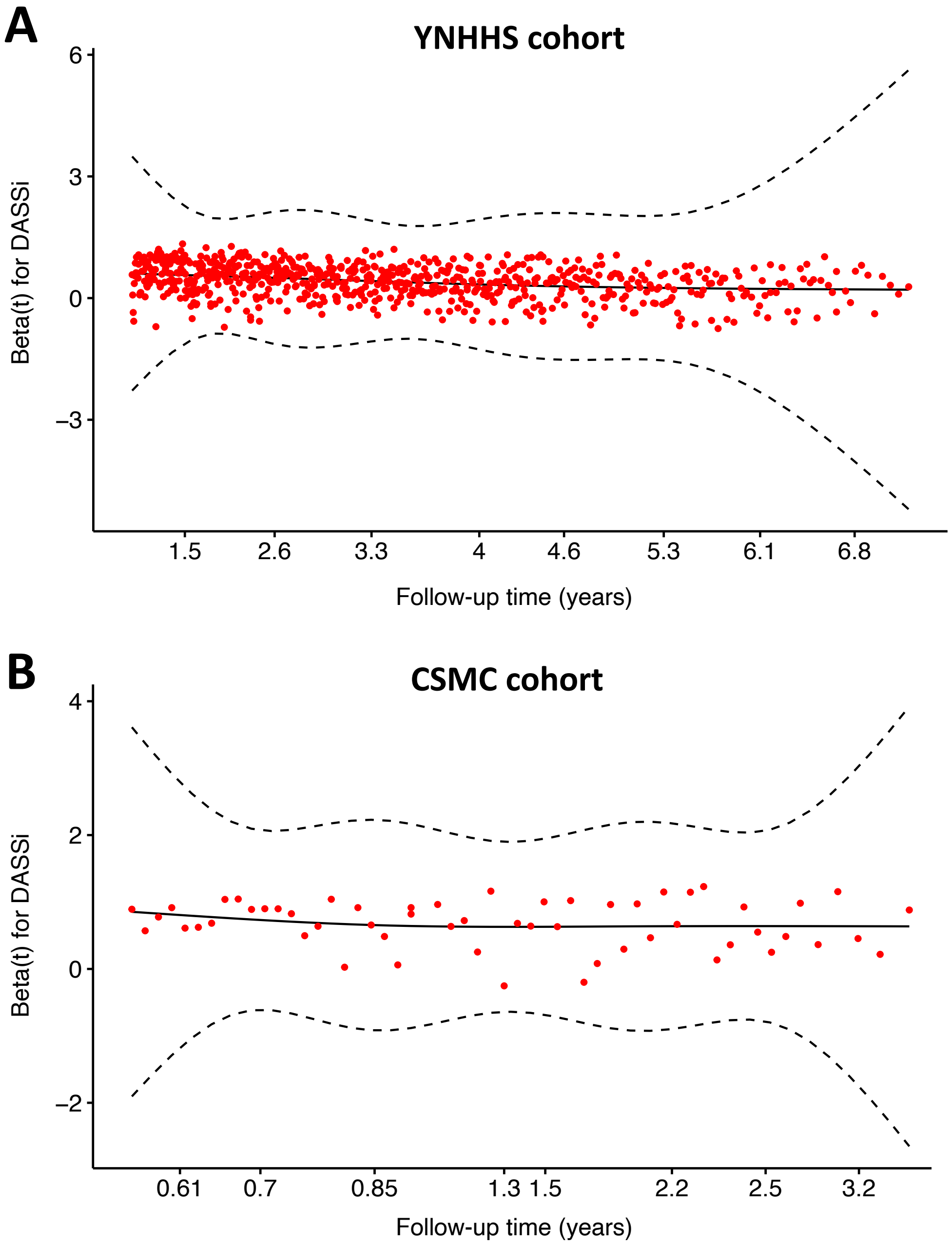


**eFigure 5 | Plots of scaled Schoenfeld residuals.** Plots providing an estimate of the time dependence of the DASSi (digital aortic stenosis index) coefficient relative to the outcome of AVR (aortic valve replacement) across **(A)** the YNHHS, and **(B)** the CSMC cohorts. A horizontal line suggests a consistent pattern across follow-up which favors the proportionality of hazards assumption. *AVR: aortic valve replacement; CSMC: Cedars-Sinai Medical Center; DASSi: digital aortic stenosis severity index; YNHHS: Yale-New Haven Health System.*

### eFigure 6

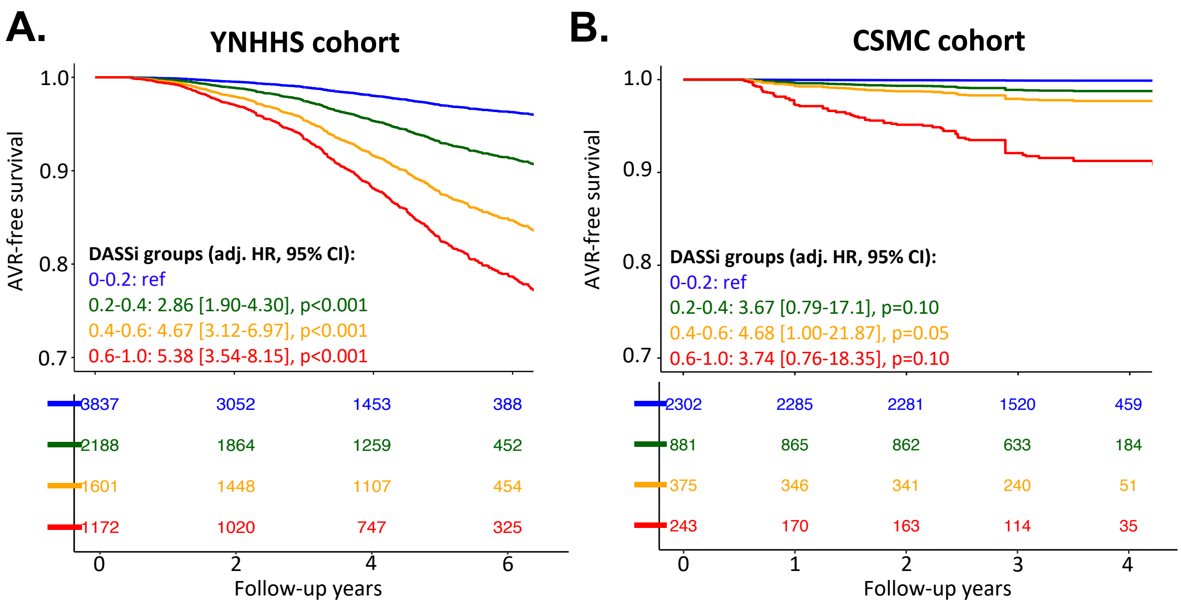


**eFigure 6 | Association of baseline echocardiographic DASSi phenotypes with future AVR.** Adjusted event-free survival curves derived from multivariable Cox regression models for aortic valve replacement (AVR) in the YNHHS **(A)** and CSMC **(B)** cohorts, respectively. Each curve represents a distinct baseline DASSi severity subgroup; associations are adjusted for age, sex, race, ethnicity, baseline peak aortic valve velocity and left ventricular ejection fraction. *AV Vmax: peak aortic valve velocity; AVR: aortic valve replacement; CI: confidence interval; CSMC: Cedars-Sinai Medical Center; DASSi: digital aortic stenosis severity index; HR: hazard ratio; LVEF: left ventricular ejection; YNHHS: Yale-New Haven Health System.*

### eFigure 7

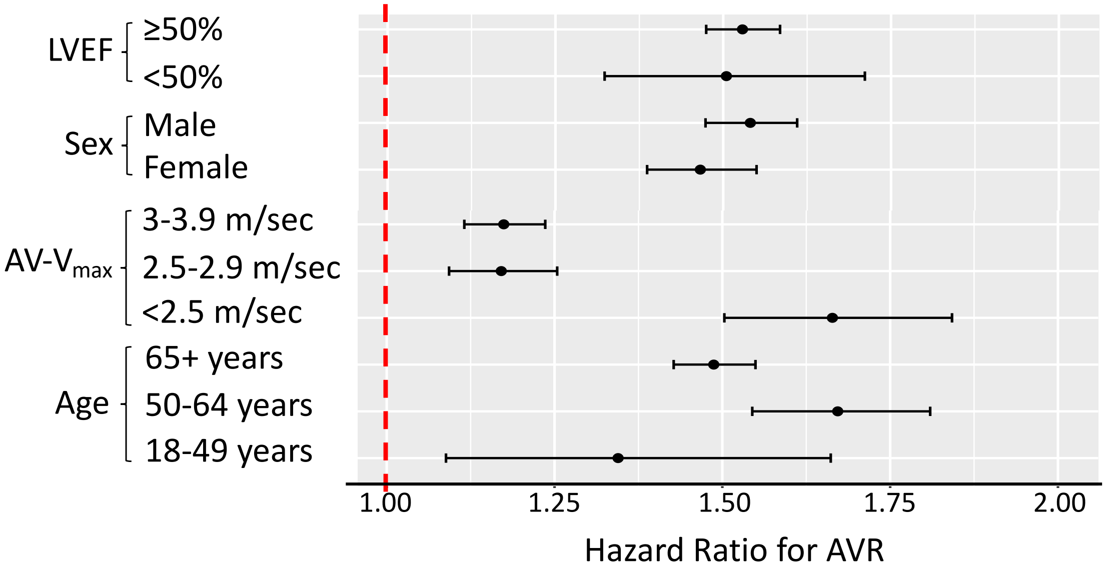


**eFigure 7 | Subgroup associations between baseline echocardiographic DASSi phenotypes and future AVR in the YNHHS cohort.** Forest plot denoting the hazard ratio and corresponding 95% confidence interval for a multivariable Cox regression of DASSi as a continuous variable (expressed in 0.1 increments) and time-to-aortic valve replacement (AVR). *AV Vmax: peak aortic valve velocity; AVR: aortic valve replacement; DASSi: digital aortic stenosis severity index; LVEF: left ventricular ejection; YNHHS: Yale-New Haven Health System.*

eFigure 8

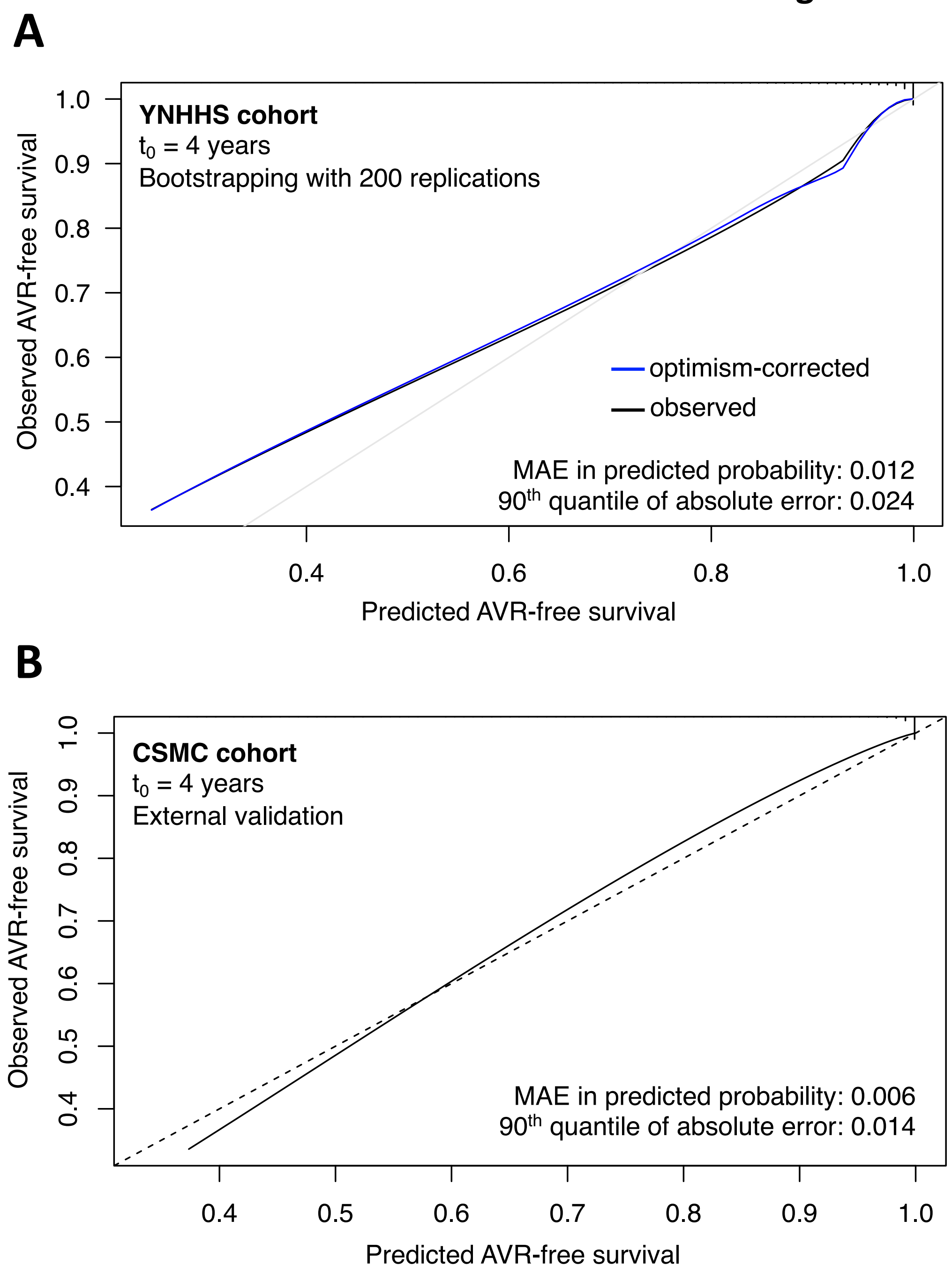


**eFigure 8 | Internal and external calibration curves for a DASSi-based model to predict future AVR.** Analyses censored at t=4 years to ensure adequate follow-up that did not exceed the length of follow-up in the CSMC cohort. **(A)** Internal calibration with cross-validation across n=200 bootstrapping replications in the YNHHS cohort. **(B)** External validation of a YNHHS-based model in the CSMC cohort. *AVR: aortic valve replacement; CSMC: Cedars-Sinai Medical Center; DASSi: digital aortic stenosis severity index; MAE: mean absolute error; YNHHS: Yale-New Haven Health System.*

**eFigure 9**

### Phenome-wide association study

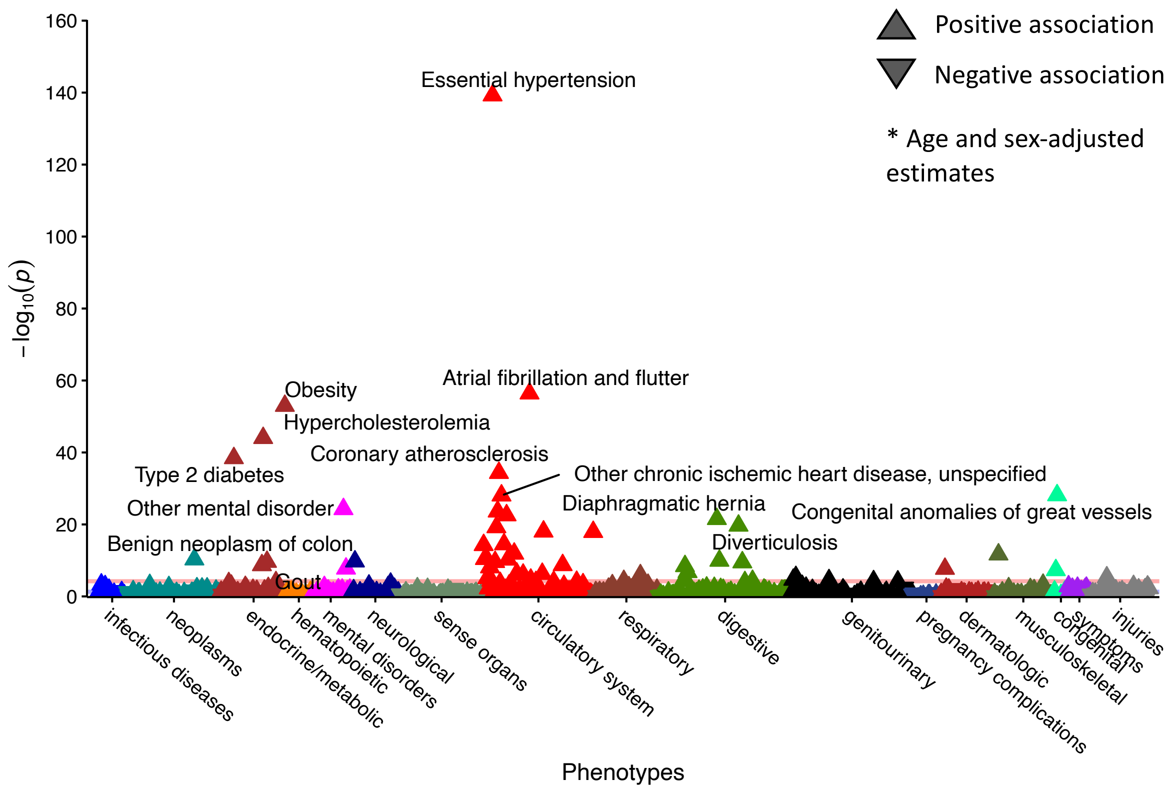


**eFigure 9 | Phenome-wide association of CMR-derived DASSi.** Phenome-wide association study of DASSi based on ICD-10 diagnosis codes and related phenotypes in the n=45,474 individuals from the UK Biobank included in the CMR analysis. The orange horizontal line denotes the significance level following Bonferroni correction. *CMR: cardiac magnetic resonance; DASSi: digital aortic stenosis severity index.*
